# An online atlas of human plasma metabolite signatures of gut microbiome composition

**DOI:** 10.1101/2021.12.23.21268179

**Authors:** Koen F. Dekkers, Sergi Sayols-Baixeras, Gabriel Baldanzi, Christoph Nowak, Ulf Hammar, Diem Nguyen, Georgios Varotsis, Louise Brunkwall, Nynne Nielsen, Aron C. Eklund, Jacob Bak Holm, H. Bjørn Nielsen, Filip Ottosson, Yi-Ting Lin, Shafqat Ahmad, Lars Lind, Johan Sundström, Gunnar Engström, J. Gustav Smith, Johan Ärnlöv, Marju Orho-Melander, Tove Fall

**Affiliations:** Department of Medical Sciences, Molecular Epidemiology and Science for Life Laboratory, Uppsala University, EpiHubben, MTC-huset, Uppsala, Sweden; Division of Family Medicine and Primary Care, Department of Neurobiology, Care Science and Society, Karolinska Institute, Huddinge, Sweden; Department of Clinical Sciences, Lund University, Malmö, Sweden; Clinical Microbiomics A/S, Copenhagen, Denmark; Department of Medical Sciences, Uppsala University, Uppsala, Sweden; Department of Medical Sciences, Clinical Epidemiology, Uppsala University, Uppsala, Sweden; The George Institute for Global Health, University of New South Wales, Sydney, Australia; The Wallenberg Laboratory/Department of Molecular and Clinical Medicine, Institute of Medicine, Gothenburg University and the Department of Cardiology, Sahlgrenska University Hospital, Gothenburg, Sweden; Department of Cardiology, Clinical Sciences, Lund University and Skåne University Hospital, Lund, Sweden; Wallenberg Center for Molecular Medicine and Lund University Diabetes Center, Lund University, Lund, Sweden; School of Health and Social Studies, Dalarna University, Falun, Sweden

## Abstract

The human gut microbiota produces a variety of small compounds, some of which enter the bloodstream and impact host health. Conversely, various exogenous nutritional and pharmaceutical compounds affect the gut microbiome composition before entering circulation. Characterization of the gut microbiota–host plasma metabolite interactions is an important step towards understanding the effects of the gut microbiota on human health. However, studies involving large and deeply phenotyped cohorts that would reveal such meaningful interactions are scarce. Here, we used deep metagenomic sequencing and ultra-high-performance liquid chromatography linked to mass spectrometry for detailed characterization of the fecal microbiota and plasma metabolome, respectively, of 8,584 participants invited at age 50 to 64 of the Swedish CArdioPulmonary bioImage Study (SCAPIS). After adjusting for multiple comparisons, we identified 1,008 associations between species alpha diversity and plasma metabolites, and 318,944 associations between specific gut metagenomic species and plasma metabolites. The gut microbiota explained up to 50% of the variance of individual plasma metabolites (mean of 4.7%). We present all results as the searchable association atlas “GUTSY” as a rich resource for mining associations, and exemplify the potential of the atlas by presenting novel associations between oral medication and the gut microbiome, and microbiota species strongly associated with levels of the uremic toxin p-cresol sulfate. The association atlas can be used as the basis for targeted studies of perturbation of specific bacteria and for identification of candidate plasma biomarkers of gut flora composition.

## Introduction

The bacteria, archaea, viruses, protozoa and fungi that reside in the gastrointestinal tract are collectively referred to as the gut microbiota. The gut microbiota is shaped by all lifetime exposures of an individual including diet, disease history, antibiotics and other medication^1^; and by intrinsic factors, such as age and host genetic variation^2^. Conversely, observational studies suggest a role of gut microbiota composition in chronic disease development e.g. cardiovascular disease, obesity, and type 2 diabetes, but evidence of causality and mechanistic understanding of these effects are largely absent^3–5^. Modification of the composition of small molecules in plasma, i.e. the plasma metabolome^3^, has been suggested as a potential mediator of gut microbiota effects on human health, as the gut microbiota produces and modifies a number of molecules, some of which are taken up into the bloodstream. Consequently, characterization of the interactions between gut microbiota and host plasma metabolites could provide crucial insights into the effects of the gut microbiota on human health.

Previous studies^6–13^ reporting associations between the gut microbiota and the circulating metabolome have been hampered by either small sample size (e.g. a few 100 samples), limited data on health-related traits, or limited resolution of gut microbiota composition (e.g. 16S rRNA sequencing) and metabolome data (e.g. NMR profiling). While these studies have shown that the gut microbiota composition is associated with at least a portion of the plasma metabolome, major questions remain. Specifically, since statistical power has been limited, moderate effect sizes or associations of rare species with metabolites have not been possible to assess. Further, there is an imminent need for a public resource of these associations as a useful tool to help the researchers better understand the gut microbiota - plasma metabolome interplay.

Here, we applied state-of-the-art high resolution deep metagenomic sequencing and mass spectrometry-based metabolite profiling to analyze samples from 8,584 individuals from SCAPIS, a well-characterized population-based study. We generated the searchable GUTSY Atlas of robust associations between the gut microbiota and host plasma metabolome including functional metabolic modules.

## Results

### Gut microbial species and plasma metabolite profiling of the SCAPIS study

SCAPIS^14^ is a prospective population-based observational study of 30,154 men and women living in six municipality regions in Sweden. A randomly selected sample of individuals aged 50 to 64 based on the population register were invited during the years 2014 to 2018 to participate in the baseline investigation. We focused on data obtained at two study sites, Malmö and Uppsala, where 9,818 fecal samples were collected at home, and from which DNA was extracted, whole-genome shotgun-sequenced, and taxonomically and functionally profiled. The taxonomical profiling resulted, at the super kingdom level, in 1,484 bacterial, 4 archaeal, 2 eukaryotic and 2 unclassified metagenomic species, from now on called species, which were identified based on their microbial gene profile, with an average of 302 species per sample (range, 26 to 579, Supplementary Table 1). In addition, 9,109 fasting venous plasma samples were collected during study site visit, which underwent metabolomics profiling using ultra-high-performance liquid chromatography linked to mass spectrometry provided data on 1,364 metabolites, of which 1,057 were annotated from 116 classes of metabolites, with an average of 1,227 measured metabolites per sample (range, 1,119 to 1,290, Supplementary Table 2). Overall, data for 8,584 individuals passed quality control for both fecal metagenomics and plasma metabolomics. The main sociodemographic and clinical characteristics of these 8,584 individuals are shown in Table 1. Further, we created a companion website (the “GUTSY Atlas”), which contains download links to all the results we generated in the current study, and allows generation of tables and figures for specific species in association with metabolites. Below, we describe some general and specific trends mined using the atlas.

**Table 1.** Main sociodemographic and clinical characteristics of the Malmö and Uppsala SCAPIS subcohorts included in the current study. Continuous variables are provided as mean (standard deviation) and categorical variables as *n* (%).

|  | **Malmö (*n* = 3,812)** | **Uppsala (*n* = 4,772)** |
| --- | --- | --- |
| Age, years | 57.4 (4.3) | 57.7 (4.4) |
| Sex: female | 2009 (52.7%) | 2451 (51.4%) |
| Place of birth |  |  |
| Scandinavia | 2977 (78.1%) | 4295 (90.0%) |
| Europe | 543 (14.2%) | 184 (3.9%) |
| Asia | 202 (5.3%) | 193 (4.0%) |
| Other | 90 (2.4%) | 100 (2.10%) |
| Body mass index, kg/m<sup>2</sup> | 27.2 (4.5) | 27.0 (4.4) |
| Systolic blood pressure, mmHg | 122 (16.4) | 125 (15.9) |
| Estimated glomerular filtration rate | 84.5 (12.1) | 87.0 (11.5) |
| Current smoker | 640 (16.8%) | 431 (9.0%) |
| Fiber intake, g/kcal<sup>1</sup> | 0.012 (0.004) | 0.012 (0.004) |
| Coffee intake |  |  |
| <2 times/d | 1116 (29.3%) | 1587 (33.3%) |
| 2–3 times/d | 1762 (46.2%) | 2217 (46.5%) |
| >4 times/d | 934 (24.5%) | 968 (20.3%) |
| Any antibiotics, last 12 months | 786 (20.6%) | 896 (18.8%) |
| Blood pressure medication<sup>2</sup> | 775 (20.3%) | 883 (18.5%) |
| Lipid-lowering medication<sup>2</sup> | 319 (8.4%) | 352 (7.4%) |
| Diabetes medication<sup>2</sup> | 170 (4.5%) | 161 (3.4%) |

<sup>1</sup>Fiber intake, adjusted for total energy intake; <sup>2</sup>Self-reported medication last 2 weeks

### Metabolite signatures of microbial diversity

We first investigated the association of microbial alpha diversity with individual plasma metabolites. Alpha diversity was estimated using the Shannon diversity index, a measure of overall microbiota richness and evenness previously reported as inversely associated with markers of metabolic health^15^. We observed that the alpha diversity was positively associated with 585, and negatively associated with 423, of the 1,364 plasma metabolites in models adjusted for age, sex, country of birth, study site and sequencing plate (Figure 1, Supplementary Table 3). Significance was based on *p*-values adjusted for multiple testing, which we report as *q*-values, using the Benjamini-Hochberg method^16^ at a 5% false discovery rate. Regarding annotated metabolites, we observed the strongest positive associations for the metabolite 5alpha-androstan-3beta,17alpha-diol disulfate (*ρ* = 0.44, *q*-value < 10^−300^), a sulfated steroid previously reported associated to alpha diversity^17^; and cinnamoylglycine (*ρ* = 0.39, *q*-value < 10^−300^), a glycine conjugate that is present in the plasma of healthy mice, but not in the plasma of germ-free mice^18^. Of the 50 metabolites with strongest positive associations with alpha diversity, 48 metabolites were associated with more than 30% of species. These observations indicate that gut microbial diversity is robustly associated with a range of specific plasma metabolites and motivated the ensuing detailed investigations of specific gut microbiota species.

**Figure 1.**
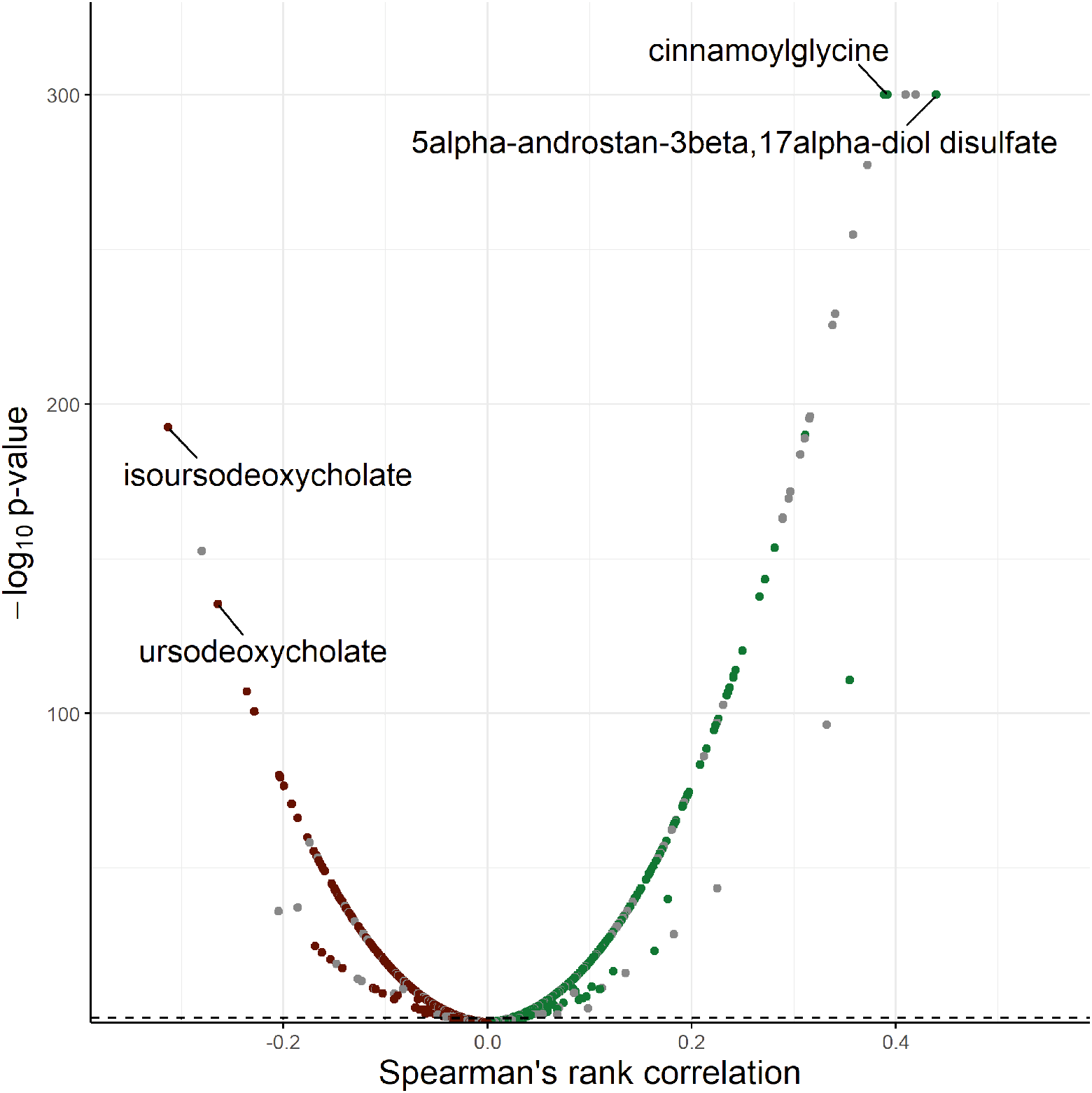
Spearman partial correlation between species alpha diversity and 1,364 plasma metabolites adjusted for age, sex, place of birth, study site and sequencing plate. The association of Shannon diversity index based on deep metagenomic sequencing of fecal samples and 1,364 plasma metabolites measured with ultra-high performance liquid chromatography linked to mass spectrometry in 8,584 participants aged 50 to 65 of the Swedish CArdioPulmonary bioImage Study. There were 585 significant positive associations and 423 significant negative associations after adjusting for multiple testing using Benjamini-Hochberg’s method at 5% false discovery rate. Green, positive associations; red, negative associations; grey, indicates the non-characterized metabolites. Labels are shown for the 2 most positively and negatively correlated characterized metabolites. The dashed line represents the multiple testing threshold. The *p*-values were capped at 10^−300^.

### Associations of gut microbiota with plasma metabolome show large variation across groups of bacteria and metabolites

We used least absolute shrinkage and selection operator (lasso) models to assess the variance in the plasma metabolome explained by the variation of the gut microbiota. We observed that the variance of 1,258 of the 1,364 metabolites was partly explained by variation in the gut microbiota (mean *r*^2^ = 4.7%, Figure 2A, Supplementary Table 4). We detected the largest variance explained (50%) for an uncharacterized common metabolite with the provisional identifier X-11850, by a combination of 306 species, with *Mogibacterium kristiansenii* contributing the most. The gut microbiota explained >30% of the variance of 13 additional metabolites, such as uremic toxin *p*-cresol sulfate (*r*^2^ = 36%, species contributing = 406, strongest contributor = *Clostridia sp*., MGS:0140) and the coffee metabolite quinate (*r*^2^ = 32%, species contributing = 464, strongest contributor = *Eubacteriales sp*., MGS:0102). For trimethylamine N-oxide, TMAO, the end-product of diet-microbiota interaction, which has been suggested involved in cardiovascular and kidney disease pathogenesis^19^, we found a rather low variance explained by the gut microbiota (*r*^2^ = 1.1%, species contributing = 55, strongest contributor = *Dialister pneumosintes*). These observations highlight the large heterogeneity in associations of gut microbiota composition with plasma metabolites.

**Figure 2.**
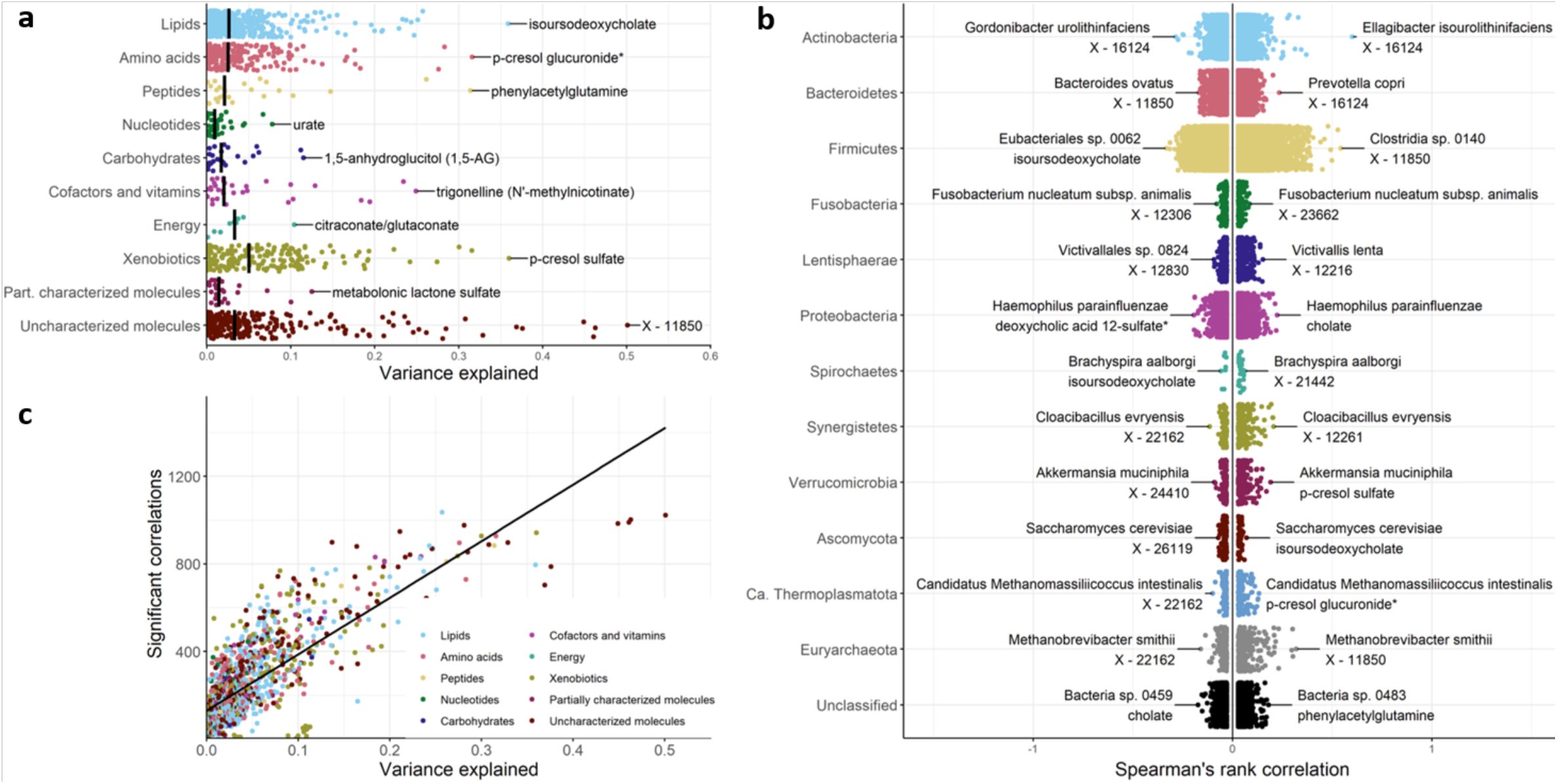
Associations of gut microbiota with plasma metabolome show great variation across groups of bacteria and metabolites. a, The variance in 1,364 plasma metabolites explained by gut microbiota in samples from 8,584 individuals aged 50 to 65 of the Swedish CArdioPulmonary bioImage Study. Models were fitted for each metabolite using least absolute shrinkage and selection operator regression using 10-fold cross-validation. The variance explained was calculated as the cross-validated *r*^2^ statistic. Metabolites were grouped by metabolic pathway and the vertical line represents the median of the variance explained for each group. The metabolite with the largest variance explained for each group is annotated. b, Partial Spearman’s rank correlations between 1,493 gut microbial species and 1,364 plasma metabolites adjusted for age, sex, place of birth, study site, Shannon diversity index and sequencing plate. Depicted are the Spearman’s rank coefficients for 168,237 significant positive associations and 150,707 significant negative associations after adjusting for multiple testing using Benjamini-Hochberg’s method at 5% false discovery rate. Associations were grouped by taxonomic phylum. c, Variance explained versus number of associated species for 1,364 plasma metabolites. Metabolites were grouped by metabolic pathway. Shown in black is the linear regression line.

### Associations of gut microbiota with plasma metabolome are many and robust over a range of lifestyle and health factors

We next assessed the links between 1,493 microbial species and 1,364 metabolites using a series of partial Spearman correlations, adjusting for age, sex, country of birth, Shannon diversity index, study site and sequencing plate. We identified significant associations (*q*-value < 0.05) in 318,944 (16%) of all tested species–metabolite pairs, of which 168,237 were in the positive direction and 150,707 in the negative (Figure 2B, Supplementary Tables 5). While all species and metabolites had at least one observed association, some species (*n* = 152) were associated with a broad range (>30%) of metabolites, with the two most common metabolites being vitamin A metabolism metabolites (>142 associations with species), and 252 metabolites were associated with a broad range (>30%) of species (Extended Figure 1). The associations between species and metabolites were not generally affected by stratification at the tertiles of body mass index (BMI), systolic blood pressure, estimated glomerular filtration rate^20^ (eGFR, a measure of kidney function), fiber intake, nor by exclusion of smokers and individuals who had been prescribed antibiotics within a year of sampling or taken medication for hypertension, dyslipidemia and/or diabetes (Pearson correlation of Spearman’s *ρ* from non-stratified vs stratified models *r* > 0.94, Extended Figure 2), findings which alleviated concerns about major confounding effects by these factors. However, we did find lower correlation of results within smokers (*r*=0.79) and individuals with cholesterol (*r*=0.79) or diabetes medication (*r*=0.55). This could either be explained by lower precision as groups were smaller (n, 332-1,074) or by effect modification by these factors for some associations. Generally, metabolites for which the variance explained by gut microbiota was high also had a high number of associations with individual species (Figure 2C). Overall, these observations show a plethora of associations between gut microbiota species and the metabolome that are in general robust over a range of lifestyle and health factors.

### Certain species are associated with multiple metabolites, often within the same class of metabolites

We observed several examples of the same microbial species having both strong positive and negative associations with metabolites within the same class of metabolites indicating that the species might affect a specific process (Figure 2B). One observed example is *Haemophilus parainfluenzae*, a bacterial species previously linked to bile tract infections^21^, that was strongly positively associated with the primary bile acid salt cholate and negatively associated with the secondary bile acid deoxycholic acid 12-sulfate. We further grouped single metabolites into metabolite classes and asked if individual species were linked to several metabolites of the same class. Overall, we found evidence supporting the enrichment for metabolite classes for 1,155 species (total 289,775 enrichments, *q*-value < 0.05, Supplementary Table 6). Among the 10 strongest enrichments, four were related to secondary bile acids, namely *Olsenella sp*. AF21-5, *Oscillospiraceae sp*. MGS:0806, *Anaerotignum faecicola* and *Peptostreptococcaceae sp*. MGS:0200. Given these findings, we set out to leverage the atlas to identify species influencing the rate of 7α-dehydroxylation forming deoxycholic acid from cholic acid, which is one of the main first steps in the formation of secondary bile acids from primary bile acids. Previous research have detected bile acid 7α-dehydroxylation activity in a limited group of bacteria such as *Clostridium species* nested within a phylogenetic tree containing members of *Blautia, Ruminococcaceae* and *Lachnospiraceae*^22^. To identify potential new species with 7α-dehydroxylation activity, we assessed gut species correlated with low plasma levels of the precursor cholic acid and increased levels of the product deoxycholic acid. Both the primary bile acid cholic acid (denoted cholate in the atlas) and the secondary bile acid deoxycholic acid (deoxycholate) had a high variance explained by the microbiota (*r*^2^ = 20% and 23%, respectively), indicating a strong impact of the variation in microbiota composition. Of the 20 species with strongest negative correlation with cholate and 20 species with strongest positive correlation with deoxycholate, seven were in common. We confirmed these findings using a model with deoxycholate/cholate ratio as outcome (all *p*-values <10^−100^). These species include novel findings indicating a role in bile acid metabolism such as *Dysosmobacter welbionis, Anaerotruncus colihominis, Clostridiales bacterium* and *Lachnospiraceae bacterium* 1 4 56FAA and two species previously linked to bile acid metabolism, *Clostridium citroniae* and *B. obeum*.

Collectively, our results indicate that certain species are associated with multiple metabolites within the same class of metabolites; this was especially prominent for secondary bile acids and their precursors.

### Functions shared by several species are linked to single metabolite abundance

Different microbiota species may share genetic elements that enable them to perform the same metabolic function. We hypothesized that such genetic elements shared by several species affect single metabolite levels. We therefore mapped microbial genes to 103 gut metabolic modules (GMM), in order to associate microbial metabolic function with species associated with single metabolites. In total, we found an enrichment of microbial functions for 1,277 plasma metabolites (total 14,305 enrichments, *q*-value < 0.05, Supplementary Table 7). Among the microbial functions with strongest enrichments, those functions encoding enzymes catalyzing the degradation of amino acids and monosaccharides such as the threonine (e.g. with 4-hydroxycoumarin), fructose (e.g. with deoxycholic acid glucuronide), sucrose (e.g. with ferulic acid 4-sulfate) and lysine degradation (e.g. with 1,5-anhydroglucitol (1,5-AG) were most prominent. Overall, these findings support that certain functions shared by several species are common in species that are associated with plasma metabolite abundances.

The above analyses clearly show the existence of specific associations between gut microbiota and the host plasma metabolome. Below, we present detailed data of the association of selected microbes and metabolites as example of information that can be mined using the GUTSY Atlas of associations of the plasma metabolome with gut microbiota. We focused on microbiota interactions with coffee metabolites, as an example of a dietary component reported to have large effects on the microbiota; with *p-*cresol sulfate, as an example of a bacteria-derived metabolite implicated in human health; and with omeprazole and metformin, medications, which are thought to have profound effects on the microbiota.

### Coffee metabolites have strong positive associations with species from the *Eubacteriales* order

Coffee is one of the most widely consumed beverages in the world and has a complex and not fully elucidated relationship with human health^23^. The PREDICT1 (*n* = 1,098) study revealed a large number of diet-microbiota associations, of which the strongest combined associations were found for coffee intake.^12^ We sought to further investigate the links between microbiota characteristics and coffee using the GUTSY Atlas data by investigating 12 established coffee metabolomic biomarkers^24,25^. We observed that 19 individual species in different combinations represented the eight most strongly associated species for each of these 12 biomarkers (as depicted in Extended Figure 3). These 19 species were all from the *Eubacteriales* order from the *Ruminococcaceae, Oscillospiraceae, Lachnospiraceae* and *Clostridiaceae* families, except *Streptococcus saliviarus*. Three species were annotated at the species level: *Clostridium phoceensis, Anaeromassilibacillus sp. Marseille-P3371* and *Streptococcus saliviarus*, which were all associated in the positive direction with all the 12 coffee biomarkers. *Clostridium phoeensis* was first identified in the gut microbiota of a healthy 28-year-old man in Marseille^26^ and has not previously been linked to any phenotypes. *Anaeromassilibacillus sp. Marseille-P3371* has been found affected by a low-protein diet in a dietary trial of chronic kidney disease patients. The commensal bacterium *Streptococcus salivarius* is one of the early bacteria colonizing the oral and gut mucosal surfaces. This species is proposed to have positive effects in the oral cavity and upper respiratory tract; it may inhibit colonization of other pathogens such as *S. pyogenes*^27^ and virulent streptococci involved in tooth decay such as *S. mutans*^28^, and also has anti-inflammatory characteristics. It is currently not known why the abundance of certain gut bacteria is positively associated with coffee intake. It does not seem to be driven by smoking behavior (Extended Figure S3), but it may be related to the metabolism of these bacteria. Of note, coffee is rich in antioxidants^29^ and affects gut motility, which could also affect the sampling and the bacterial community^30^. In summary, we report novel association of previously reported coffee biomarkers with the abundance in the gut microflora with a set of bacteria from the *Eubacteriales* order and with *Streptococcus saliviarus*.

### *Faecalibacterium prausnitzii* is negatively associated with the uremic toxin *p*-cresol and phenylacetylglutamine

In the current study, we observed that 36% of the variation in *p*-cresol sulfate plasma levels was explained by the variation in gut microbiota — one of the highest proportions of explained variation of all metabolites. The bacterial metabolite *p*-cresol is classified as a uremic toxin and is produced during bacterial tyrosine fermentation in the large intestine and accumulated in patients with kidney failure, and its levels are associated with worse outcomes.^31,32^ In a germ-free mouse model of chronic kidney disease, transplantation of fresh microbiota from end-stage renal disease patients led to an increase in serum levels of *p*-cresol sulfate and other uremic toxins^33^. This was interpreted to mean that the aberrant gut microbiota in renal patients aggravates the disease by modulating uremic toxin levels, and highlights the importance of better characterization of the uremic toxin-producing microbiota. We found that *p*-cresol sulfate and the related metabolite *p*-cresol glucuronide as well as the glutamine-derived phenylacetylglutamine^34^ showed much stronger associations with several species from the *Eubacteriales* order, including novel positive associations with *Intestinimonas massiliensis* (*p-*value = 3.7 × 10^−244^), than other established and proposed uremic toxins, such as hippurate, indoxyl sulfate, TMAO, and 3-carboxy-4-methyl-5-propyl-2-furanpropanoic acid (Extended Figure 4). This association supports that the *Eubacteriales* order, formerly called *Clostridiales*, is one of the most prolific phenol compound-generating bacterial subgroups that produce *p*-cresol sulfate as a tyrosine fermentation end product^35^. Importantly, we also found several strains of the *Faecalibacterium prausnitzii* strongly inversely associated with *p-*cresol levels and phenylacetylglutamine. Interestingly, *F. prausnitzii* was one of the depleted species in the microbiota of renal patients, compared to healthy controls^33^, and its reduced levels have been linked to more severe stages of renal disease.^36^ We performed additional models stratified by eGFR, and found slightly stronger associations in the individuals with lower kidney function (Extended Figure 4). In summary, we identify a number of species that are positively or negatively associated with *p*-cresol and phenylacetylglutamine levels, which sets the foundation for future studies into perturbation of the gut flora in chronic kidney failure to reduce uremic toxins.

### The microbiota of omeprazole users is characterized by the increased abundance of oral bacteria and is enriched for bacterial functions related to carbohydrate metabolism

Omeprazole is a selective proton pump inhibitor (PPI) commonly used for treatment of acid-related upper gastroduodenal diseases, and sold over-the-counter. In the current study, we observed strong positive associations between presence of omeprazole in plasma and bacteria belonging to *Veillonella* species (e.g., *V. parvula, V. dispar* and *V. atypica*) and *Streptococcus* species (e.g., *S. anginosus, S. oralis subsp oralis, S gordonii, S. salivarius, S. parasanguinis* and *S. mutans*), all parts of the normal oral microbiota. This is in line with findings from a recent studies^13,37^ that reported that PPI use was associated with an increased abundance of several taxa common to the oral flora, such as *Veillonella* and several *Streptococcus* species. Interestingly, *V. parvula* is reported to have a mutualistic relationship with *S. mutans* by co-aggregating and transforming metabolic products of other carbohydrate-fermenting bacteria^38^. We also observed that the diabetes-associated microbiota-derived metabolite imidazole propionate showed a similar pattern of bacterial associations^39^. With regards to the potential function of omeprazole-associated bacteria, we found that functional modules linked to degradation of lactose, galactose, fructose, trehalose, pentose phosphate, sucrose and the Entner-Doudoroff pathway (substrate: glucose) were strongly enriched (all *p*-values <10^−5^) for bacterial species positively associated with omeprazole, pointing again to carbohydrate-fermentation. Although we only investigated omeprazole and no other types of PPI, earlier studies have demonstrated similar effects of different PPI types on the gut microbiota^13,37^. Taken together, the current study provides strong support for the notion that PPI use is associated with consistent alteration of gut microbiota, characterized by the increased abundance of bacteria common in the oral flora with an enrichment for bacterial functions related to carbohydrate metabolism.

### Metformin and gut microbiota

Metformin is a widely used anti-diabetic drug that has been associated with profound changes in the gut microbiota composition, and also with gastrointestinal side effects such as bloating and discomfort^40,41^. Here we identified 152 species, whose abundances were associated with metformin, of which an increased abundance of *Escherichia marmotae* and *E. coli*, and decreased abundance of *Romboutsia timonensis* and *Intestinibacter bartlettii* were the strongest associations. These top findings are in accordance with earlier studies reporting a significant enrichment of *E. coli* in the gut microbiota of metformin users^13,40,41^ and a decreased abundance in *R. timonensis* and *I. bartletii*^13^, as well as with a recent randomized trial that showed that metformin treatment in overweight/obese individuals results in an increased abundance of *E. coli* and a decreased abundance of *I. bartlettii* at 6 and 12 months of metformin treatment^42^. Further, an increase of *Ruminococcus torques* was reported at both time points in that study^42^ which is also supported by an earlier study^13^ and our study (*p*-value = 7.4 × 10^−7^). *R. timonensis* is a new species that was recently isolated from the human gut^43^ and has not been associated with use of metformin prior to the Mueller et al. study^42^. In species associated with metformin, we found an enrichment for bacterial functional modules related to sugar metabolism and sugar transport systems. Interestingly, the top four microbial functions most strongly associated with metformin were involved in sugar metabolism, such as the third strongest association overall in the atlas, with the pentose phosphate pathway (oxidative phase, *p*-value = 2.6 × 10^−14^) the Entner-Doudoroff pathway and fructose degradation. Taken together, our results confirm and expand previous findings that metformin treatment is associated with profound changes of the gut microbiota composition, and that bacteria carrying genes enabling carbohydrate metabolism are in higher abundance in metformin users.

## Discussion

We performed the largest and most detailed association study of the gut microbiota and host plasma metabolites to date and present the results as the online GUTSY Atlas, which can be used as the starting point for targeted studies of perturbation of specific bacteria and to identify candidate plasma biomarkers of gut flora composition. The analysis revealed 318,944 associations of individual microbial species with metabolites and confirm and substantially expand previous studies in the area^6–12^. This resource is non-targeted and thereby encompass large parts of the described and undescribed human gut microbial community and the plasma metabolome, enabling researchers with varying interests to benefit from the data.

We observe a large variation in the association of gut microbiota species with plasma metabolites, where certain metabolites such as *p*-cresol and secondary bile acids having strong associations with multiple bacterial species, and others, such as nucleotides showing few associations. We report that certain species are associated with multiple metabolites within the same class of metabolites; this was especially prominent for secondary bile acids and their precursors, secondary bile acids are produced by the gut bacteria and involved in fat and oil digestion^44^.

We also detect a number of novel observations in terms of specific biomarkers, metabolites and drugs, such as the association of coffee biomarkers with a set of bacteria from the *Eubacteriales* order and with *Streptococcus saliviarus*, which is regarded as a competitor to more pathogenic strains of *Streptococci*. These results support previous findings that coffee intake, one of the most consumed beverages globally, have effects on the composition of the gut microbiota, which warrants further investigations in the possible links with health.

We find a number of species strongly associated with *p*-cresol levels. *P*-cresol is regarded as an important uremic toxin, produced during bacterial tyrosine fermentation in the large intestine and accumulated in patients with kidney failure causing further damage to the kidney, and can only be marginally removed by dialysis.^31,32^ We identify several substrains of *F. prausnitzii* with variable association with *p*-cresol, indicating that certain strains may have larger effects than others. The results from the GUTSY Atlas could hence be used as a foundation for designing future studies of gut flora perturbation in chronic kidney failure to reduce uremic toxins with the purpose to decrease kidney disease progression.

Our study provides strong support for the notion that PPI use is associated with consistent alteration of gut microbiota, characterized by the increased abundance of bacteria common in the oral flora with an enrichment for bacterial functions related to carbohydrate metabolism. PPI use is common in the population and found associated with a number of traits in observational studies, such as small intestine bacterial overgrowth. The health impact of PPI-related changes of the gut microbiota warrants further investigation.

Our findings also confirm many of those reported in the previously largest high-resolution gut microbiome – plasma metabolome study from the TwinsUK adult twin registry (*n* = 859)^8^. For example, for the top ten associations annotated to the species level in the TwinsUK, nine were available in our study of which seven were replicated, i.e., associations between *F. prausnitzii* and *p*-cresol sulfate, *p*-cresol glucuronide, phenylacetylglutamine and deoxycholate, *Methanobrevibacter smithii* and threonate, *Roseburia inulinivorans* and *p*-cresol sulfate, and *E. coli* and phenylacetylglutamine. Interestingly, their top finding of a strong association of *Barnesiella intestinihominis* with plasma levels of sebacate (decanedioate) was not replicated in our study, although both the species and the metabolite were present in our data. Further, in the current study, the gut microbiota explained 50% of the variance in the uncharacterized molecule X-11850 and explained >30% of the variance in other metabolites, such as coffee metabolite quinate (*r*^2^ = 0.32) and uremic toxin *p*-cresol sulfate (*r*^2^ = 0.36). This aligns with the recent study by Bar et al.^10^, which analyzed these associations using similar methods but in two smaller Israeli cohorts (*n* = 491, replication in 1,004 participants from TwinsUK and 245 from IMI-DIRECT)^10^. For example, in the study by Bar et al., X-11850 exhibited the second highest variance explained (*r*^2^ = 0.49), and quinate (*r*^2^ = 0.45) and *p*-cresol sulfate (*r*^2^ = 0.41) were also among the top 10 strongest associations. The identified large overlaps with previous studies and the current study indicates that findings are in general robust over different populations and analytical platforms. Given the improved statistical power in the current study, we expanded the number of findings from 254 associations of the TwinsUK study to 318,944 associations in the GUTSY Atlas, also including associations of moderate effect size and for more rare species. We also confirm and expand previous findings that metformin treatment is associated with profound changes of the gut microbiota composition, and that bacteria carrying genes enabling carbohydrate metabolism are in higher abundance in metformin users, in line with data from an intervention study^41^. These results were among the strongest in the overall enrichment analysis of all drugs, such as for pentose phosphate pathway and PTS transport modules.

### Strengths and limitations

The major strengths of the current study are the sample size, high-resolution data and the easy-to-use companion website. While we replicated here the findings of other studies, confirming the quality of the generated data, we also identified many novel associations between oral medication and the gut microbiome, and microbiota species strongly associated with levels of the uremic toxin *p*-cresol sulfate. The cohort analyzed in the current study is more than three times bigger than the previously largest study in which gut microbiota were analyzed by 16S sequencing and associated with NMR-based plasma metabolome profiling (*n* = 2,309)^7^ and 10 times bigger than that of a previously largest study in which gut microbiota were analyzed by high-resolution metagenomics and associated with mass spectrometry-based plasma metabolome profiling (*n* = 859)^8^, which allowed us to assess also associations of moderate size and with rare metabolites and species. Another strength is the deep phenotyping of the SCAPIS study, which allowed detailed sensitivity analyses of potential confounders and effect modifiers^14^. Consequently, the presented association atlas is based on a large well-characterized sample and state-of-the art analytical methods for microbiota and metabolomics which will enable well-powered *in silico* exploration of the potential metabolic effect of various bacteria of interest and for identifying candidate plasma biomarkers of gut flora composition.

However, some limitations of the current study should be recognized. First, the study population comprises mostly Scandinavian-born individuals of predominantly European descent. While the top findings for this cohort were similar to those of samples for the UK and Israel, generalizations to populations on other continents might not be valid. Second, the observational nature of the cross-sectional study design makes residual confounding a potential issue and causal inference difficult. Nonetheless, for food-derived metabolites, such as the coffee metabolite quinate, and drugs, such as omeprazole and metformin, the causal direction from the medication/food intake to the gut microbiota is most likely, although it could still be confounded by factors that co-vary with the food and medication type. Conversely, for metabolites produced by the gut microbiota, such as secondary bile acids, we assume the causal direction from the gut microbiota to the plasma metabolome. Any causal links should, however, be verified in the future by using experiments or causal inference methods, such as Mendelian randomization^45,46^. Third, similar to previous studies, the associations were analyzed using ranked-based non-parametric models, which hinders the interpretation of actual effect sizes. Fourth, although the number of annotated metabolites and species is high in this study, many of the identified associations were between unknown metabolites and species for which no reference genome is currently available. This makes putting the novel findings in context challenging. We plan to update the companion website as additional metabolites and species are characterized in the future.

In summary, we here identified a vast number of robust associations between the gut microbiota and the plasma metabolome, and report these to the community in the GUTSY Atlas, a comprehensive online resource for an interactive investigation of the associations. These findings add to the knowledge of the vast interactions of the gut microbiota and human metabolism and will generate insights into human biology and identification of potential novel biomarkers of gut flora composition. We anticipate that the GUTSY Atlas will be of immense benefit for the scientific community, reducing the need for collecting and analyzing their own samples.

## Data Availability

Restrictions apply to the availability of individual level data, which can only be used with ethical approval and, hence, are not publicly available. However, the data are available from the authors upon reasonable request and with a written permission from the Swedish Ethical Review Authority and the SCAPIS Data Access Board.

## Methods

### Study sample

SCAPIS is a prospective population-based study of 30,154 men and women, aged 50–65 years, living in six municipality regions in Sweden. It was designed with the main aim to improve risk prediction and understanding of cardiovascular disease, chronic pulmonary obstructive disease, and related metabolic disorders^14^. After a pilot study in 2012, recruitment was initiated in 2014 and completed in 2018. Individuals were randomly recruited from the population register, with a participation rate of 50%. The present study is based on the data for 11,285 participants from the test centers in Malmö (*n* = 6,251) and Uppsala (*n* = 5,034). Of these, fecal metagenomics data were available for 9,818 individuals (Malmö, *n* = 4,980; Uppsala, *n* = 4,838) and plasma metabolomics data were available for 9,109 individuals (Malmö, *n* = 4,126; Uppsala, *n* = 4,983). After exclusion of 2,879 participants who lacked complete metagenomics and metabolomics dataset or whose samples failed quality control, 8,584 individuals were included as the sample in the current study.

All study participants provided a signed informed consent at the first site visit. The study adheres to the Declaration of Helsinki and was approved by the Swedish Ethics Review Authority (Etikprövningsmyndigheten Dnr 2010-228-31M, Dnr 2018-315).

### Gut microbiome sample collection and preprocessing

Participants received a pre-packaged fecal sample collection kit (barcoded tubes, gloves, Ziploc bags, and a paper collection bowl) including instructions on how to collect the sample at home. The participants were asked to store the samples at –20 °C in the home freezer until the study site visit. Once received in the laboratory, the samples were kept at –20 °C for 0–7 d until transport to the central biobank for storage at –80 °C. Finally, the samples were shipped on dry ice to Clinical Microbiomics A/S (Copenhagen, Denmark) for metagenomics analysis. Samples were analyzed in a random order and 20 samples were analyzed in replicate.

Fecal DNA was extracted using NucleoSpin® 96 Soil kit (740787; Macherey-Nagel; Germany). Negative and positive controls were included. Samples were subjected to 5 min of bead beating at 2,200 rpm, with 1.2 μg of DNA obtained on average per sample. The sequencing of metagenomes was performed using an Illumina Novaseq 6000 system (Illumina, USA). On average, 26.3 million read pairs (7.9 Gb) were generated per sample for Malmö samples, and 25.3 million read pairs (7.6 Gbp) were generated per sample for Uppsala samples. Sequencing reads with adapters, containing >10% ambiguous bases, those with >50% bases with Phred quality score <5, and reads mapped to the human reference genome GRCh38 were removed using Bowtie 2 v.02.3.2^47^ with default settings. The remaining reads were assembled using MEGAHIT v. 1.1.1^48^ and mapped using BWA mem v. 0.7.16a^49^ to a newly created gene catalog of 14 M non-redundant microbial genes from three main sources: data from the current study, data from Pasolli et al.^50^, and 3,488 publicly available genomes of isolated microbial strains relevant to the human gut microbiome^51–53^. Metagenomic species were defined as co-abundant gene groups from the gene catalog that fulfilled previously established quality criteria^54^. In each species, 100 highly correlated and distinct signature genes were identified and used for abundance profiling. Overall, 1,985 species were identified in the gene catalog. The number of gene counts for every sample mapped to the signature genes of a species determined the count number of that species. A minimum of 3 read pairs had to be mapped to the species signature genes for it to be considered as detected. Species relative abundances were calculated by dividing the counts of each species by the effective length of its signature genes, and then normalizing each sample to sum to 100%. The species relative abundances were (natural) log+1 transformed, and species with at least 100 non-zero values were included in further analysis. After quality control, 1,493 species were included, with an average of 302 per sample. Shannon diversity was calculated using the *vegan* R package^55^. For taxonomic annotation, catalog genes were compared to those in the NCBI RefSeq^56^ database (downloaded on 2 May 2021). Species-level taxonomy was assigned to metagenomic species with >75% of genes annotated to a single species. For the genus, family, order, class, and phylum annotations, different thresholds were used (60, 50, 40, 30, and 25%, respectively).

For functional annotation, catalog genes were annotated to the gut metabolic modules^57^ (GMM) database using EggNOG-mapper^58^ v 2.0.1. Potential functional profiles were determined for species that contained at least 2/3 of the enzymes/protein genes needed for the functionality of a particular GMM module. If an alternative reaction pathway within a module existed, only one such reaction pathway was required. All reaction pathways were required for modules with fewer than four steps.

### Plasma metabolome sample collection and preprocessing

Venous blood samples were collected from the participants during the study site visit after an overnight fast. The samples were stored at –80 °C in the biobank until shipping to Metabolon Inc. for plasma metabolome analysis (Durham, NC, USA)^59^. Samples were handled and analyzed in random order together with different quality control standards, namely, pure water, solvents used for metabolite extraction, a pool of human plasma samples maintained by Metabolon Inc., and a pool of study participants’ samples. Proteins were removed by methanol precipitation with vigorous shaking using Glen Mills GenoGrinder 2000 and centrifugation. To maximize metabolite identification, four processes were used in parallel: reverse phase (RP)/ultrahigh performance liquid chromatography–tandem mass spectroscopy (UPLC-MS/MS) with negative-ion mode electrospray ionization (ESI), hydrophilic interaction chromatography (HILIC)/UPLC-MS/MS, and two separate RP/UPLC-MS/MS resolutions with positive-ion mode ESI. Peak identification and quantification, and quality control were performed using Metabolon’s hardware and software. For each metabolite, for each instrument plate (144 samples), the peak measurement areas were divided by the median peak area of samples in that batch. Metabolite measurements that failed to reach the detection threshold were imputed from the minimum observed value for that metabolite. Metabolites were annotated by matching to Metabolon’s library of more than 3,300 purified standards and unknown compounds based on the retention time-index, mass-to-charge ratio, and chromatography data. As part of the annotation process, two types of metabolic pathways were assigned to each metabolite: (1) “metabolite class”, which includes broad metabolic pathway terms, and (2) “metabolite subclass”, which includes narrow metabolic pathway terms. Metabolites with at least 100 values above the detection threshold were included in the present study. Metabolites other than drug metabolites were (natural) log+1 transformed. Drug metabolites were converted to binary values (present or absent). Overall, 1,364 metabolites that passed quality control were included in the analyses.

### Statistical analysis

Analyses were performed and plots were created with R (version 4.1.1, https://cran.r-project.org/). Partial Spearman correlations were calculated for Shannon diversity index and each metabolite using the *ppcor*^60^ R package. Correlation estimates were adjusted for age, sex, place of birth, study site (Uppsala/Malmö), and metagenomic sequencing plate. Association *p*-values were adjusted for multiple testing using the Benjamini–Hochberg method at 5% false discovery rate (FDR). Similarly, for each species, partial spearman correlations were calculated for each metabolite and adjusted for age, sex, place of birth, study site (Uppsala/Malmö), Shannon diversity index, and metagenomic sequencing plate. Sensitivity analyses were performed by stratification of the data by potential mediators and confounders using the same model.

Lasso models were used to estimate the amount of variance of each metabolite explained by a combination of species using the *glmnet*^61^ R package. A model with the lambda that gave the minimum mean 10-fold cross-validated error was chosen and *r*^2^ statistics were calculated based on cross-validated errors.

Enrichment analysis of ranked association *p*-values was performed using the *fgsea*^62^ package for positive and negative Spearman’s rank coefficients separately as one-sided test. The enrichment *p*-values for positive and negative coefficients were combined and adjusted for multiple testing using the Benjamini–Hochberg method at 5% FDR. Enrichment analysis was done using GMM modules and metabolite subclasses.

## Code availability

Code related to the analyses in this study are available at https://github.com/MolEpicUU/GUTSY_Atlas.

## Companion website

A companion website to the article, containing download links to all the summary statistics data generated for the current study and further study-related searchable material will be made accessible once the article has been published.

## Acknowledgements

We acknowledge the financial support from the European Research Council [ERC-2018-STG801965 (TF); ERC-CoG-2014-649021 (MO-M); ERC-STG-2015-679242 (JGS)], the Swedish Research Council [VR 2019-01471 (TF), 2018-02784 (MO-M), 2019-01015 (JÄ), 2020-00243 (JÄ), 2019-01236 (GE), 2021-02273 (JGS)]; the Swedish Heart-Lung Foundation [Hjärt-Lungfonden, 20190505 (TF), 20200711 (MO-M), 20180343 (JÄ)], 20200173 (GE); 20190526 (JGS)]; and Formas [2020-00989 (SA)]. The main funding body of the Swedish CArdioPulmonary bioImage Study (SCAPIS) is the Swedish Heart-Lung Foundation. The study is also funded by the Knut and Alice Wallenberg Foundation; the Swedish Research Council, and VINNOVA (Sweden’s Innovation agency); the University of Gothenburg and Sahlgrenska University Hospital; Karolinska Institutet and Stockholm county council; Linköping University and University Hospital; Lund University and Skåne University Hospital; Umeå University and University Hospital; and Uppsala University and University Hospital.

The computations and data handling were made possible by resources from project sens2019512 provided by the Swedish National Infrastructure for Computing (SNIC) at Uppsala Multidisciplinary Center for Advanced Computational Science (UPPMAX), partially funded by the Swedish Research Council through grant agreement no. 2018-05973.

## Author contributions

G.E, G.S, J.Ä., MO-M. and T.F. obtained the funding. K.F.D., S.S-B., U.H., J.Ä., MO-M. and T.F. designed the study and developed the concept. L.B, G.E, J.G.S, J.S., J.Ä., MO-M. and T.F. collected the data. N.N, A.E, J.B.K, H.B.N performed metagenomic analysis and bioinformatics, K.F.D performed all association analysis and created the web atlas, MO-M. and T.F. coordinated the study. K.F.D, G.B, C.N, D.N., J.Ä, MO-M. and T.F. wrote the manuscript. All authors provided interpretation of the results and critical feedback on the manuscript.

## Competing interests

N.N., A.E. and J.B.H., and H.B.N. are employees of Clinical Microbiomics. The funders had no role in study design, data collection and analysis, decision to publish or preparation of the manuscript. J.Ä. has served on advisory boards for AstraZeneca and Boehringer Ingelheim and have received lecturing fees from AstraZeneca and Novartis, all unrelated to the present project. The remaining authors declare no competing interests.

## Corresponding author

Correspondence and requests for materials should be addressed to Tove Fall.

Supplementary Information is available for this paper.

**Extended Figure 1.**
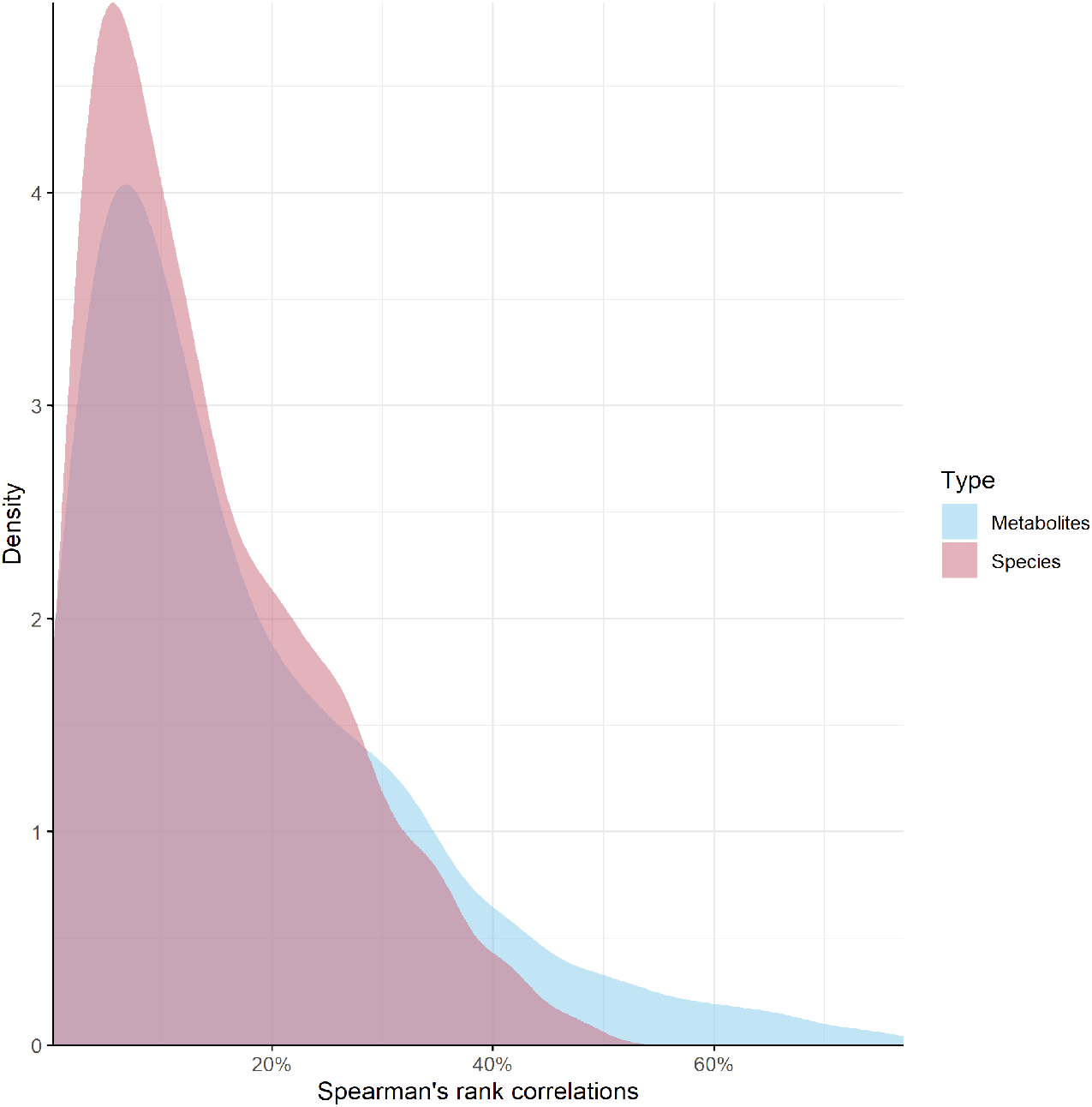
Density plot of significant Spearman’s rank correlations for each microbial species (red) and plasma metabolite (blue) as a percentage of total species (*n* = 1,493) and metabolites (*n* = 1,364), respectively. Based on partial Spearman’s rank correlations between 1,493 gut microbial species and 1,364 plasma metabolites adjusted for age, sex, place of birth, study site, Shannon diversity index and sequencing plate.

**Extended Figure 2.**
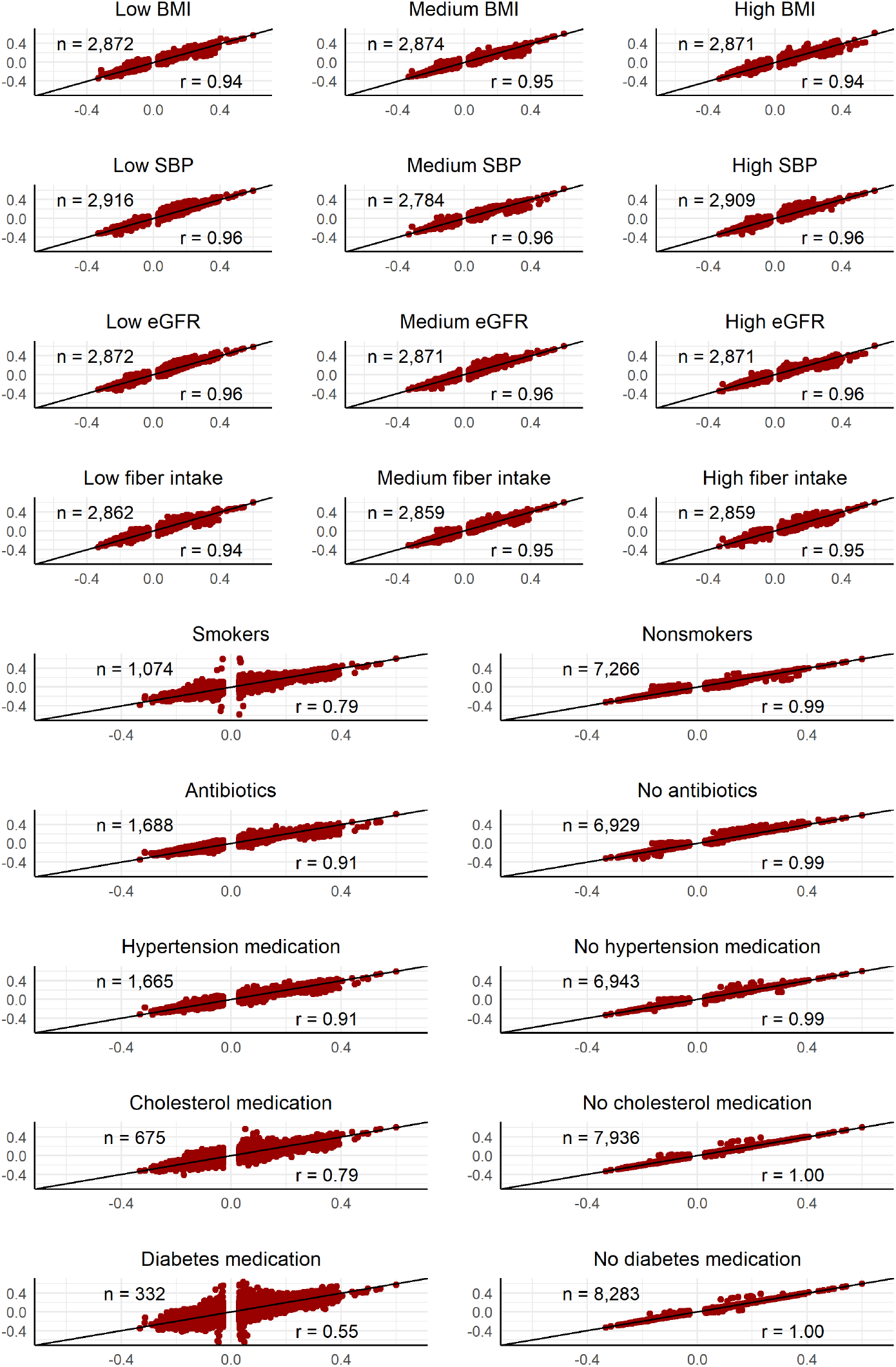
Model for all individuals compared to models stratified for potential confounders and mediators for 318,944 significant associations. X-axis, partial Spearman’s rank coefficients for the base model of 8,584 individuals; y-axis, partial Spearman’s rank coefficients for models stratified for potential confounders and mediators. Specific considerations: BMI, body mass index; SBP, systolic blood pressure; Fiber intake, adjusted for energy intake; Antibiotics, antibiotics prescribed in the year before sampling; eGFR, estimated glomerular filtration rate calculated with the CKD-EPI Study equation^20^; Hypertension, cholesterol and diabetes medication, self-reported medication last 2 weeks.

**Extended Figure 3.**
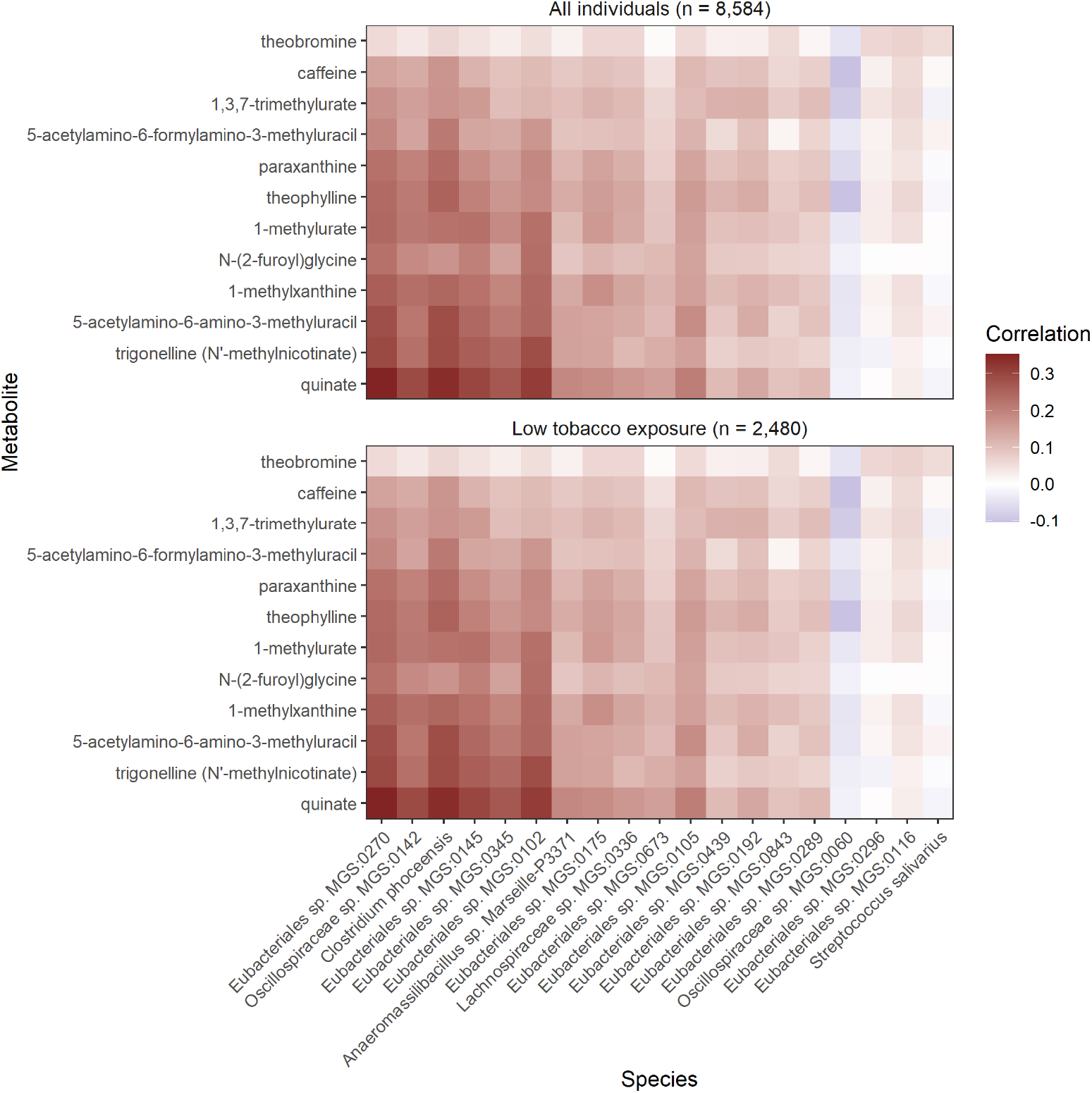
Previously reported coffee-associated biomarkers and their association with microbial species in the full dataset (n=8,584) and in 2,480 individuals who had low tobacco exposure (never smoked, no snus, no other nicotine products, no smokers in house and no smokers in the work environment). Partial Spearman’s rank correlations were calculated for 1,493 gut microbial species and 1,364 plasma metabolites adjusted for age, sex, place of birth, study site, Shannon diversity index and sequencing plate. Coffee biomarkers were selected based on studies by Shi *et al*.^24^ and Rothwell *et al*.^25^ For each coffee-associated metabolite, the 8 strongest associations with microbial species were selected based on their *p*-value, and the unique subset of those 19 species is shown.

**Extended Figure 4.**
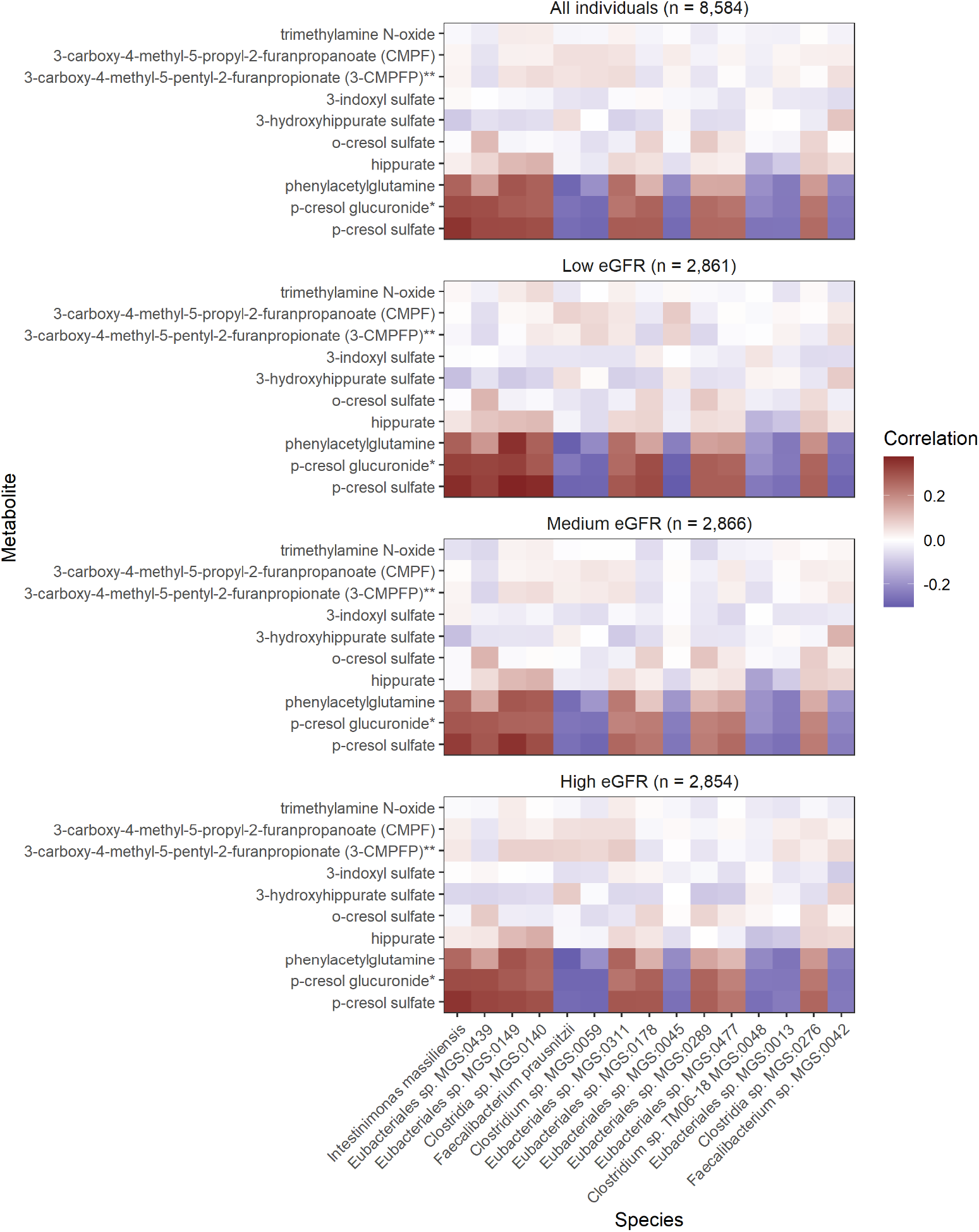
Spearman’s partial rank correlation coefficients for *p*-cresol sulfate and other uremic toxins with the 15 microbial species most strongly correlated (based on *p*-value) with *p*-cresol sulfate, and stratified by kidney function assessed by estimated glomerular filtration rate, eGFR. The associations were assessed using partial Spearman’s rank correlation between 1,493 gut microbial species and 10 candidate uremic toxins adjusted for age, sex, place of birth, study site, Shannon diversity index and sequencing plate. Specific considerations: * and ** denotes metabolites annotated without an internal standard; eGFR were estimated with the CKD-EPI Study equation^20^.

## Notes

### Author Declarations

The study adheres to the Declaration of Helsinki and was approved by the Swedish Ethics Review Authority (Etikprövningsmyndigheten Dnr 2010-228-31M, Dnr 2018-315).

